# Genetic, Clinical, and Sociodemographic Profile in Individuals with Diagnosis or Family History of Hypertrophic Cardiomyopathy: Insights from a Prospective Cohort

**DOI:** 10.1101/2025.01.24.25321106

**Authors:** Emerson de Santana Santos, Gabriel da Costa Kuhn, Antônio Guilherme Cunha de Almeida, João Victor Andrade Pimentel, Newton Vital Figueiredo Neto, Bráulio Cruz Melo, Daniel Pio de Oliveira, Luiz Flávio Galvão Gonçalves, Ana Beatriz Leite Aragão, Bárbara Letícia Lima dos Santos, Beatriz Carolina de Araujo Pereira, Beatriz Luduvice Soares, Caio da Silva Ferreira, Donizete Ferreira de Sousa Junior, João Paulo Dias Costa, Júlia Maria Teixeira Barros, Júlia Souza Diniz, Larissa Rebeca da Silva Tavares, Laís Prado Smith Lima, Luana Dias Xavier, Pedro Lucas Cardozo Barros, Silvia Sayonara Silveira Campos, Vinícius Barbosa dos Santos Sales, Willian Moreira Leão e Silva, Yussef Sab, Enaldo Vieira de Melo, Irlaneide da Silva Tavares, Antônio Carlos Sobral Sousa, Joselina Luzia Menezes Oliveira

**Author notes:** Author to whom correspondence should be addressed. **CORRESPONDING AUTHOR** Emerson de Santana Santos. Cláudio Batista St., Palestina, Aracaju, Sergipe, 49060-676, University Hospital. (SANTOS, E. S.; ALMEIDA, A. G. C.; PIMENTEL, J. V. A.; GONÇALVES, L. F. G.; ARAGÃO, A. B. L.; PEREIRA, B. C. A.; SOARES, B. L.; FERREIRA, C. S.; SOUSA JUNIOR, D. F.; COSTA, J. P. D.; BARROS, J. M. T.; DINIZ, J. S.; LIMA, L. P. S.; XAVIER, L. D.; BARROS, P. L. C.; CAMPOS, S. S. S.; SALES, V. B. S.; SILVA, W. M. L.; SAB, Y.; MELO, E. V.; SOUSA, A. C. S.; OLIVEIRA, J. L. M.). (OLIVEIRA, D. P.). (KUHN, G. D. C.; NETO, N. V. F.; TAVARES, L. R. S.). (SANTOS, B. L. L.). (MELO, B. C.; OLIVEIRA, D. P.).

## Abstract

**Background:** Hypertrophic Cardiomyopathy (HCM) is a genetic cardiac disorder characterized by left ventricular hypertrophy without secondary causes. Diagnosis relies on imaging techniques, such as echocardiography or cardiac magnetic resonance imaging (MRI), which show a maximal end-diastolic wall thickness ≥15 mm in adults. Wall thicknesses of 13– 14 mm may also support the diagnosis in individuals with a family history of HCM or sudden cardiac death (SCD), factors that strongly predict positive genetic test results and guide genetic testing decisions. However, current guidelines advise against routine genetic testing in phenotype-negative relatives unless there is a confirmed genetic diagnosis in the proband or testing is directed by a cardiovascular genetics expert. This study analyzes the genetic, clinical, and epidemiological profiles of individuals with HCM (positive phenotype) or asymptomatic patients with a family history of SCD or unconfirmed HCM.

**Methods:** We analyzed HCM patients, as defined by the 2024 AHA/ACC/AMSSM/HRS/PACES/SCMR guideline, and asymptomatic individuals with a family history of SCD or unconfirmed HCM, performing genetic testing with a 19-gene panel to assess pathogenic mutations and implications for family screening and management.

Subjects with HCM, as defined by the 2024 AHA/ACC/AMSSM/HRS/PACES/SCMR guidelines, or those with a negative phenotype but a positive family history of sudden cardiac death (SCD) or unconfirmed HCM, underwent genetic testing using a 19-gene panel associated with HCM and its phenocopies.

**Results:** Among 200 participants (58% male, median age 52 years), pathogenic/probably pathogenic variants were identified in 62 (31% – [95% CI: 24.7 to 37.9]), classified as “positive genotype,” 81 (40.5%) had variants of uncertain significance (VUS), and 57 (28.5%) had negative results. Of the total, 130 (65%) met ACC/AHA clinical criteria for HCM, while 70 (35%) had only a family history of SCD or unconfirmed HCM. Positive genotype rates were 37.7% in HCM-diagnosed individuals and 18.6% in those with a negative phenotype but positive family history (p = 0.006). Among positive genotypes, 77.4% involved sarcomeric genes (primarily MYH7 and MYBPC3, 79.2%), and 22.6% involved phenocopy-related genes, predominantly TTR (92.9%), linked to cardiac amyloidosis. Family history of SCD was more frequent in positive genotypes (68%) than negative genotypes (46%, p = 0.004). Median interventricular septal thickness was 17 mm in positive genotypes and 15 mm in negative genotypes (p < 0.001).

**Conclusion/Discussion:** The frequency of pathogenic variants in sarcomeric genes aligns with existing literature, but TTR-related cardiac amyloidosis was notably higher, suggesting greater prevalence in this population. Interventricular septal thickness >17 mm and a family history of SCD were strong predictors of positive genetic tests. While genetic testing should be guided by family screening and counseling, a family history of SCD or unconfirmed HCM may justify testing, especially when a pathogenic variant is identified in a family member.

## INTRODUCTION

Hypertrophic Cardiomyopathy (HCM) is a relatively common hereditary cardiac condition with an estimated prevalence of 1 in 500 individuals (0.2%) in the general population. It exhibits significant phenotypic and genetic variability, as well as a heterogeneous clinical course, affecting both sexes and diverse ethnic, cultural, and racial groups. Factors such as consanguinity and genetic diversity influence its prevalence and clinical expression, particularly in underrepresented populations [1,2].

The 2024 AHA/ACC/AMSSM/HRS/PACES/SCMR guidelines define HCM as left ventricular hypertrophy (LVH) not attributable to cardiac, systemic, or metabolic diseases. This encompasses cases with identified sarcomeric variants as well as those of indeterminate genetic etiology [3]. Histologically, HCM is marked by myocyte hypertrophy, disarray, and interstitial fibrosis, leading to diastolic dysfunction, ventricular arrhythmias, and an increased risk of sudden cardiac death (SCD), particularly in younger individuals. Diagnosis is primarily based on echocardiography or cardiac magnetic resonance imaging (MRI), the latter being the gold standard for detailed assessment. A wall thicknesses of 13–14 mm may also support diagnosis in individuals with a family history or a positive genetic test [3,4,5].

Advances in genetics since the 1990s have identified HCM’s monogenic inheritance pattern. Approximately 70% of cases result from mutations in sarcomeric genes such as MYBPC3 and MYH7, while other sarcomeric genes (e.g., TNNI3, TNNT2, TPM1) account for 1–5%. Non-sarcomeric gene variants, including ACTN2, ALPK3, and CSRP3, are rarer but relevant to its etiology. These discoveries have facilitated precision medicine approaches [6,7].

Next-generation sequencing (NGS) and expanded gene panels have improved diagnostic precision and differentiation of HCM from phenocopies like Fabry disease (GLA) and transthyretin cardiac amyloidosis (TTR). Despite overlapping phenotypic features, these conditions have distinct etiologies, prognoses, and treatments, underscoring the importance of accurate differential diagnosis [8].

Clinically, HCM is highly variable, ranging from asymptomatic cases to those with dyspnea, chest pain, or syncope. While it can manifest at any age, it is often diagnosed in adolescence or early adulthood. Disease progression is associated with complications such as arrhythmias and SCD [4,9]. Management options include pharmacological therapy, septal myectomy, alcohol septal ablation, and implantable cardioverter defibrillators (ICDs), which enhance quality of life and reduce the risk of heart failure and SCD [10].

Recent studies highlight the clinical and genetic differences in HCM across ethnic groups. In Brazil, increased mortality rates have been reported in the Northeast and Southeast regions, predominantly affecting white men over 40 years. Researchers advocate for early diagnosis protocols and effective therapeutic strategies to reduce mortality and improve quality of life [11].

Genetic testing plays a crucial role in etiological diagnosis, family screening, and distinguishing HCM from other causes of LVH. The CLINGEN consortium has identified eight genes with definitive associations to HCM, emphasizing the importance of these tests in clinical management and emerging therapies, such as gene-based treatments. For relatives of individuals with pathogenic variants, predictive testing and regular clinical surveillance, including ECG and echocardiography, are recommended under current guidelines [12,13].

This study aims to analyze the genetic, clinical, and epidemiological profile of patients with a prior diagnosis of hypertrophic cardiomyopathy (positive phenotype) and asymptomatic individuals with a family history of sudden death and/or unconfirmed HCM.

## MATERIALS AND METHODS

### Study Design

This is a prospective, cross-sectional study conducted between June 2021 and August 2024. The study involved non-random, consecutive samples of patients with a previous diagnosis of HCM, referred by cardiologists. The research adhered to ethical standards and was approved by the Research Ethics Committee at the Federal University of Sergipe (CAAE 50634021.0.0000.5546, opinion: 5.793.007).

### Inclusion Criteria

Adults aged 18 years or older with a confirmed diagnosis of HCM based on echocardiography and/or cardiac magnetic resonance imaging (CMR), following the diagnostic criteria established by the American Heart Association (AHA), a family history of HCM, or a family history of unconfirmed HCM and/or sudden cardiac death (SCD).

### Exclusion Criteria

Individuals with hypertrophy attributable to other known causes such as coronary artery disease, valvopathies, hypertensive cardiomyopathy, or dilated cardiomyopathy.

### HCM Diagnosis Definition

The diagnosis of HCM was established by transthoracic echocardiography (TTE) based on specific criteria, such as maximum ventricular hypertrophy (LVH) unexplained by other causes, ≥15 mm in any segment of the left ventricle or ≥13 mm in individuals with a family history of HCM. Obstructive presentations were defined by a gradient ≥30 mmHg in the left ventricular outflow tract, either at rest, after Valsalva maneuver, or in orthostasis.

### Molecular Testing

Genetic confirmation of HCM diagnosis was defined as the primary outcome, i.e., positive phenotype/genotype. Individuals with pathogenic or likely pathogenic variants in the genes tested were classified as having a positive genotype. Genes analyzed included ACTC1, FLNC, LAMP2, MYL2, PRKAG2, TNNI3, TTR, CSRP3, GLA, MYBPC3, MYL3, PTPN11, TNNT2, DES, JPH2, MYH7, PLN, TNNC1, and TPM.

### Sample Characterization Variables

Sociodemographic variables included gender, age, origin, income, and type of healthcare assistance. Clinical variables included comorbidities such as diabetes, hypertension, and dyslipidemia. Additionally, the study recorded data on hypertrophic ventricular wall thickness (in millimeters) and clinical manifestations, including acroparesthesias, angiokeratomas, anhidrosis, arrhythmias, psychiatric disorders (anxiety and depression). Other clinical signs, such as elevated serum creatinine, corneal verticillata, hypoacusis, heart failure, renal insufficiency (microalbuminuria, proteinuria), and gastrointestinal disorders.

### Statistical Analysis

Statistical analysis was performed using R software (Version 2024.09.0, Build 375). Categorical variables were expressed as absolute numbers and percentages, with 95% confidence intervals. The Chi-squared or Fisher’s exact tests were applied for categorical variables. Continuous variables were expressed as mean ± standard error, and distributions were analyzed with the Kolmogorov-Smirnov test. For non-parametric distributions, the Mann-Whitney U test was used to compare groups.

### Ethical and Legal Principles

This study followed the ethical principles of the Declaration of Helsinki and was approved by the Research Ethics Committee at the Federal University of Sergipe. Participation was voluntary, with all participants being informed about the risks and benefits of the study. Participants provided written informed consent (Termo de Consentimento Livre e Esclarecido - TCLE), ensuring compliance with ethical and legal requirements.

## RESULTS

In this study, 200 patients from 152 distinct families were analyzed. Among them, 130 individuals (65% [95% C.I.: 57.9–71.6]) fully met the diagnostic criteria for Hypertrophic Cardiomyopathy (HCM) established by the American Heart Association (AHA). The remaining 70 participants had a family history of unconfirmed HCM or sudden cardiac death and also underwent genetic testing. The sample consisted of 115 males (58%), with a mean age of 53 years (standard deviation: 17.0). Of the participants, 77 (39%) were dependent on the Brazilian public healthcare system.

The most prevalent comorbidities were systemic arterial hypertension (61%), diabetes mellitus (20%), and dyslipidemia (58%). Additionally, 5.1% of the patients had an implantable cardioverter defibrillator, and 28% were classified as obese to varying degrees. Regarding pharmacological treatment, the most frequently prescribed medications included beta-blockers (68%), statins (59%), and angiotensin-converting enzyme inhibitors/angiotensin receptor blockers (ACEIs/ARBs), prescribed to 52% of the participants.

### Genetic Analysis

Among the sample, 62 patients (31% [95% C.I.: 24.7–37.9]) tested positive for variants classified as pathogenic or likely pathogenic (P/LP) through genetic analysis, whereas 138 (69% [95% C.I.: 62.1–75.3]) were genotype-negative. Within the genotype-positive subgroup, 77.4% (95% C.I.: 65.0–87.1) exhibited genetic alterations in sarcomeric genes, while 22.6% (95% C.I.: 12.9–35.0) harbored variants in genes associated with phenocopies. Among genotype-negative patients, 89 distinct variants of uncertain significance (VUS) were identified in 81 individuals, accounting for 58.7% (95% C.I.: 50.0–67.0) of this group.

The most frequent genes in our study were MYH7, observed in 29 out of 62 cases (46.77% [95% C.I.: 34.35% – 59.19%]), TTR in 13 cases (20.97% [95% C.I.: 10.83% – 31.10%]), and MYBPC3 in 9 cases (14.52% [95% C.I.: 5.75% – 23.28%]). A list of all genes is available in Table 1 in the supplementary materials.

**Table 1.** Genetic Variants Identified in the Study Population. The table lists the genetic variants identified in the study, each row details the affected gene, ACMG classification, nucleotide change, and amino acid change.

| FAMILY | VARIANT | ACMG<br>classification | NUCLEOTI<br>DE<br>CHANGE | AMINO ACID<br>CHANGE |
| --- | --- | --- | --- | --- |
| **CX** | CSRP3 | Likely Pathogenic | 536C>T | Thr179Met |
| **XX** | CSRP3 | Pathogenic | 715G>A | Asp239Asn |
| **CXIX** | MYBPC3 | Pathogenic | 3662del | Leu1221Argfs*16 |
| **CXXX** | MYBPC3 | Pathogenic | 3662del | Leu1221Argfs*16 |
| **CXXX** | MYBPC3 | Pathogenic | 3662del | Leu1221Argfs*16 |
| **I** | MYBPC3 | Pathogenic | 1484G>A | Arg495Gln |
| **I** | MYBPC3 | Pathogenic | 1484G>A | Arg495Gln |
| **LI** | MYBPC3 | Pathogenic | 1800del | Lys600Asnfs*2 |
| **LI** | MYBPC3 | Likely Pathogenic | 1800del | Lys600Asnfs*2 |
| **XLV** | MYBPC3 | Pathogenic | Arg453Cys | Leu1221Argfs*16 |
| **XXVIII** | MYBPC3 | Pathogenic | 3662del | Leu1221Argfs*16 |
| **CIII** | MYH7 | Pathogenic | 2389G>A | Ala797Thr |
| **CIII** | MYH7 | Pathogenic | 2389G>A | Ala797Thr |
| **CXII** | MYH7 | Pathogenic | 1357C>T | Arg453Cys |
| **CXX** | MYH7 | Pathogenic | c.715G>A | Asp239Asn |
| **CXXI** | MYH7 | Pathogenic | 1357C>T | Arg453Cys |
| **CXXXI** | MYH7 | Pathogenic | 2167C>G | Arg723Gly |
| **LIX** | MYH7 | Pathogenic | 1357C>T | Arg453Cys |
| **LV** | MYH7 | Pathogenic | 715G>A | Asp239Asn |
| **LVII** | MYH7 | Pathogenic | 715G>A | Asp239Asn |
| **LVII** | MYH7 | Pathogenic | 1750G>C | Gly584Arg |
| **LVIII** | MYH7 | Pathogenic | 715G>A;p | Asp239Asn |
| **LXI** | MYH7 | Pathogenic | 746G>A | Arg249Gln |
| **LXXV** | MYH7 | Likely Pathogenic | 2389G>A | Ala797Thr |
| **LXXV** | MYH7 | Likely Pathogenic | 2389G>A | Ala797Thr |
| **LXXV** | MYH7 | Pathogenic | 2389G>A | Ala797Thr |
| **LXXXIV** | MYH7 | Likely Pathogenic | 2389G>A | Ala797Thr |
| **XCIX** | MYH7 | Pathogenic | 2012G>A | Arg671His |
| **XI** | MYH7 | Pathogenic | 1357C>T | Arg453Cys |
| **XI** | MYH7 | Pathogenic | 1357C>T | Arg453Cys |
| XI | MYH7 | Pathogenic | 1357C>T | Arg453Cys |
| XII | MYH7 | Pathogenic | 1357C>T | Arg453Cys |
| XII | MYH7 | Pathogenic | 1357C>T | Arg453Cys |
| XIX | MYH7 | Pathogenic | 1750G>C | Gly584Arg |
| XLIX | MYH7 | Pathogenic | 2389G>A | Ala797Thr |
| XLVI | MYH7 | Pathogenic | 1750G>C | Gly584Arg |
| XVII | MYH7 | Pathogenic | 1750G>C | Gly584Arg |
| XVII | MYH7 | Pathogenic | 1750G>C | Gly584Arg |
| XXXV | MYH7 | Pathogenic | 715G>A | Asp239Asn |
| XXXVII | MYH7 | Pathogenic | 2389G>A | Ala797Thr |
| III | PTPN11 | Pathogenic | 836A>G | Tyr279Cys |
| CIX | TNNI3 | Pathogenic | 575G>A | Arg192His |
| XVIII | TNNI3 | Pathogenic | 470C>T | Ala157Val |
| XVIII | TNNI3 | Pathogenic | 470C>T | Ala157Val |
| XVIII | TNNI3 | Pathogenic | 470C>T | Ala157Val |
| CXLI | TNNT2 | Pathogenic | 856C>T | Arg286Cys |
| CXXVIII | TNNT2 | Pathogenic | 418C>T | Arg140Cys |
| CXXXIX | TNNT2 | Pathogenic | 877C>T | Arg293Cys |
| LII | TNNT2 | Likely Pathogenic | 877C>T | Arg293Cys |
| CLI | TTR | Pathogenic | 424G>A | Val142Ile |
| CXXVII | TTR | Pathogenic | 424G>A | Val142Ile |
| L | TTR | Pathogenic | 424G>A | Val142Ile |
| LVI | TTR | Pathogenic | 424G>A | Val142Ile |
| LXIII | TTR | Pathogenic | 424G>A | Val142Ile |
| LXXIX | TTR | Pathogenic | 424G>A | Val142Ile |
| LXXXIII | TTR | Pathogenic | 424G>A | Val142Ile |
| XCII | TTR | Pathogenic | 424G>A | Val142Ile |
| XIV | TTR | Pathogenic | 424G>A | Val142Ile |
| XLVIII | TTR | Pathogenic | 424G>A | Val142Ile |
| XLVIII | TTR | Pathogenic | 424G>A | Val142Ile |

| FAMILY | VARIANT | ACMG classification | NUCLEOTIDE CHANGE | AMINO ACID CHANGE |
| --- | --- | --- | --- | --- |
| XXV | TTR | Pathogenic | 424G>A | Val142Ile |
| XXV | TTR | Likely Pathogenic | 5330-2A>G | Splice acceptor |

When stratified by gene, the subgroup with pathogenic/likely pathogenic variants in the TTR gene had a median age of 77 years (IQR: 66–79), significantly higher than the subgroup with sarcomeric gene variants, which had a median age of 45 years (IQR: 35–59, p < 0.001).

**Figure 1.**
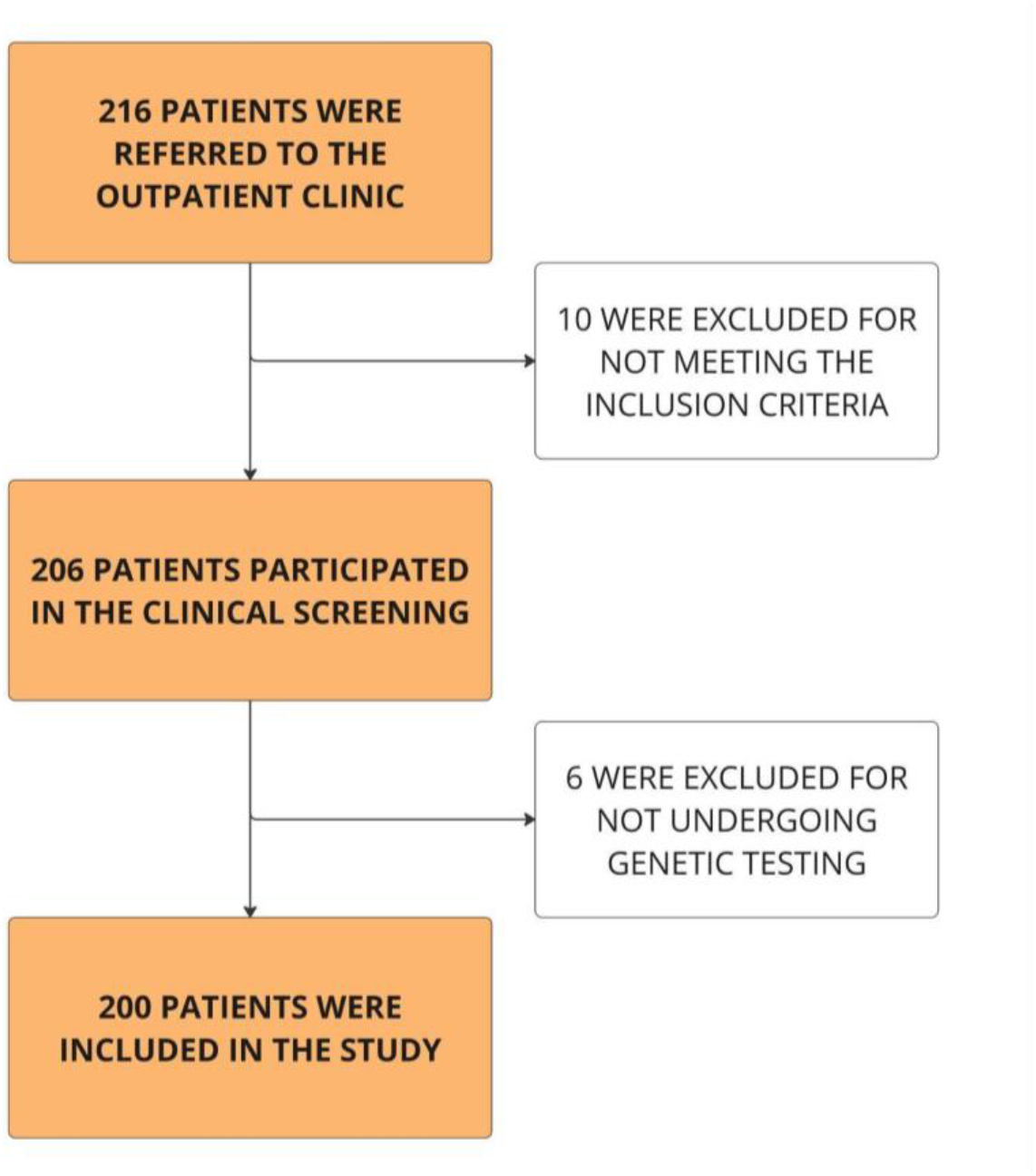
Sample Flowchart. The flowchart illustrates the sample size at each stage and outlines the reasons for exclusion from the study.

**Figure 2.**
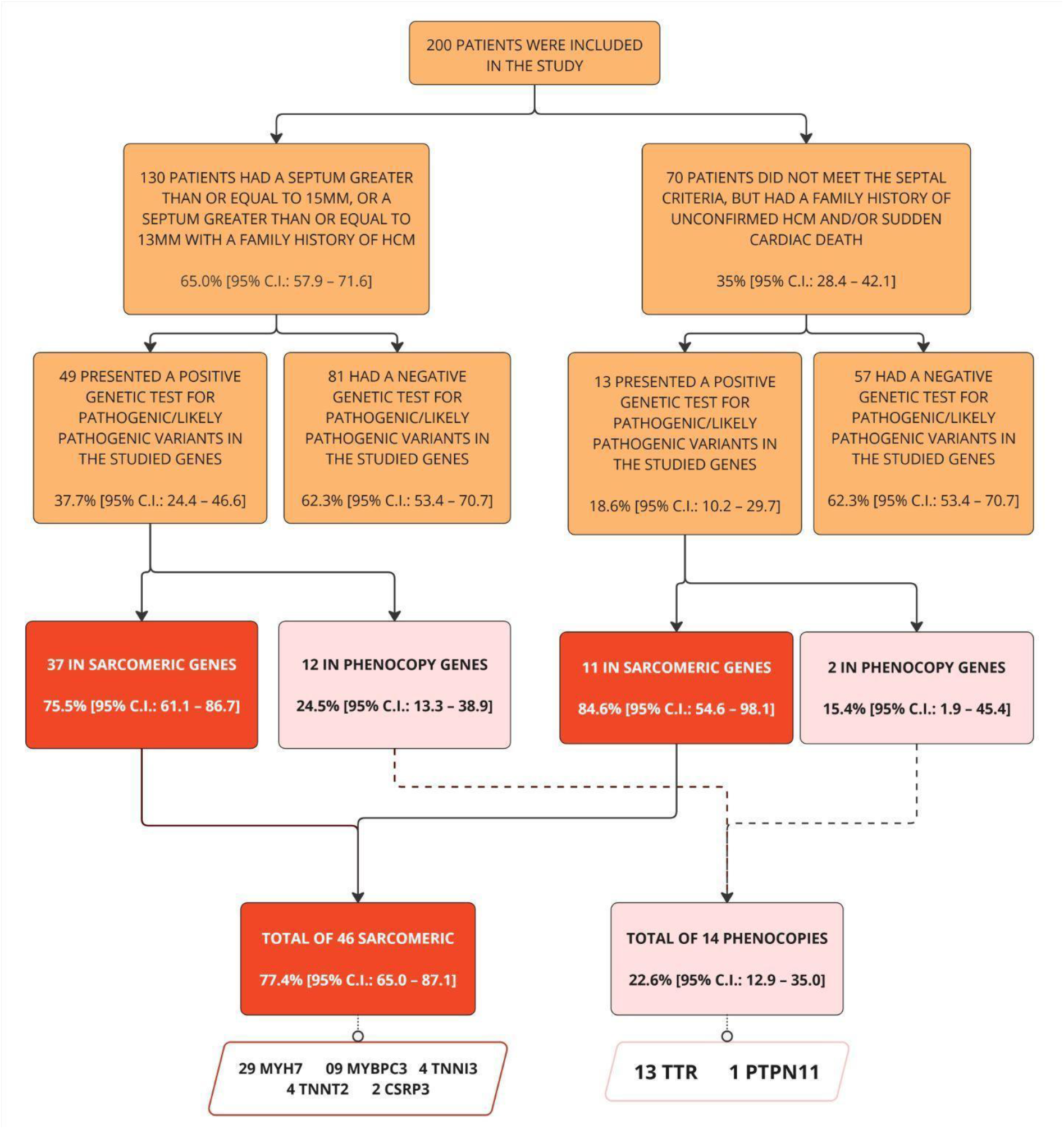
Flowchart of Criteria and Genetic Diagnosis. The flowchart depicts the subdivision created within the sample and highlights the genetic diagnoses across subgroups and the total sample.

The median interventricular septal thickness was 14.1 mm (IQR: 11.0–17.0) on echocardiography and 16.0 mm (IQR: 13.0–20.0) on cardiac magnetic resonance imaging, with an overall median of 15.0 mm (IQR: 13.0–18.0) when considering either diagnostic modality. The analysis of these diagnostic approaches demonstrated comparable results, as depicted in Figure 3.

**Figure 3.**
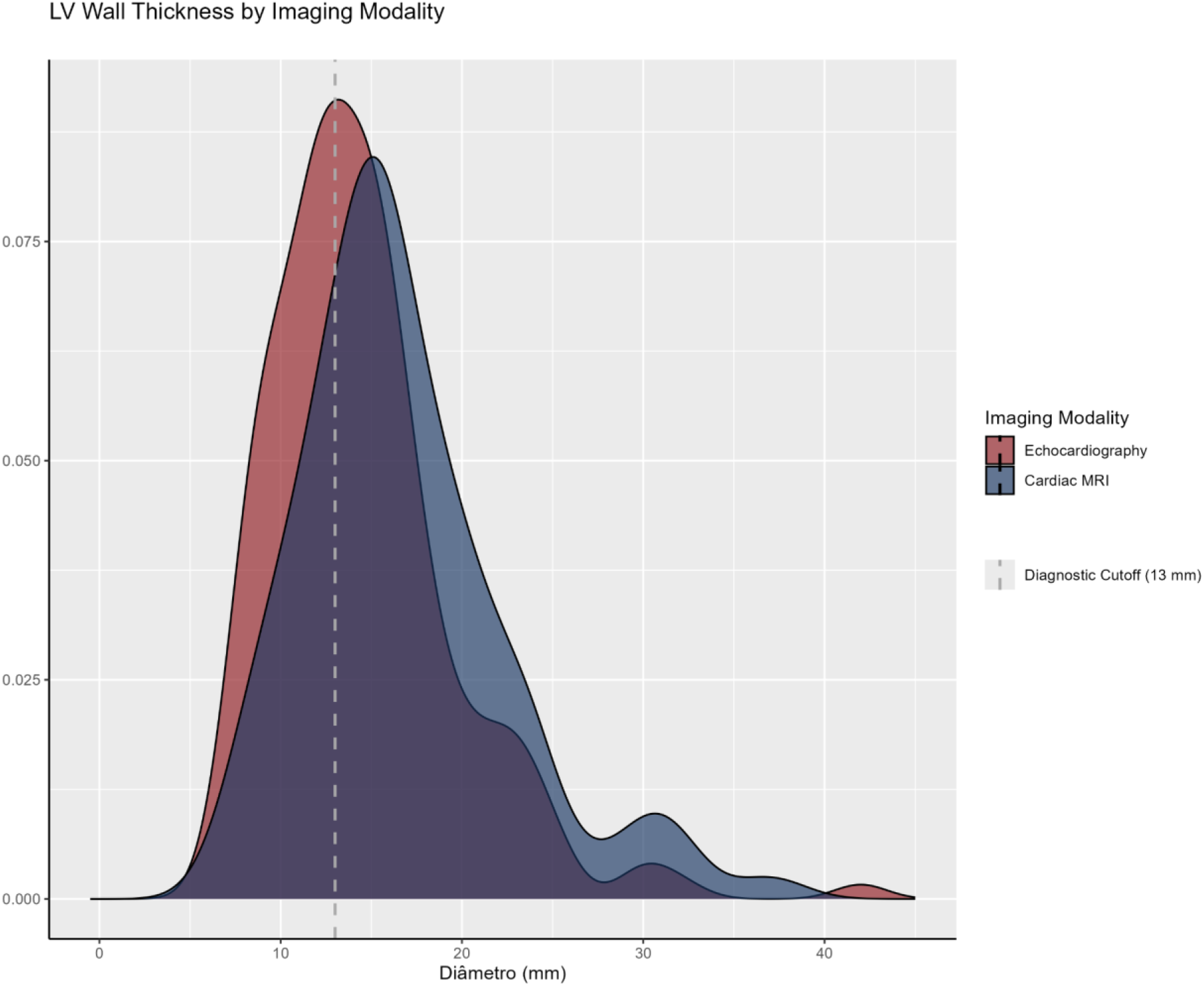
LV Wall Thickness by Imaging Modality. Shows the distribution of left ventricular wall thickness (measured in millimeters) by imaging modality: Echocardiogram (Red) and Cardiac MRI (Blue). The dashed vertical line at 13mm represents the minimal diagnostic cutoff for HCM, consistent with the 2024 AHA/ACC/AMSSM/HRS/PACES/SCMR guideline.

When stratified by genetic diagnosis, patients with a positive genotype exhibited a greater mean interventricular septal thickness (17.7 mm vs. 15.0 mm; p < 0.001). The median septal thickness among patients with VUS and those without pathogenic/likely pathogenic variants was 14.4 mm and 16.0 mm, respectively. Figure 4 illustrates the distribution of septal thickness across genetic diagnosis subgroups.

**Figure 4.**
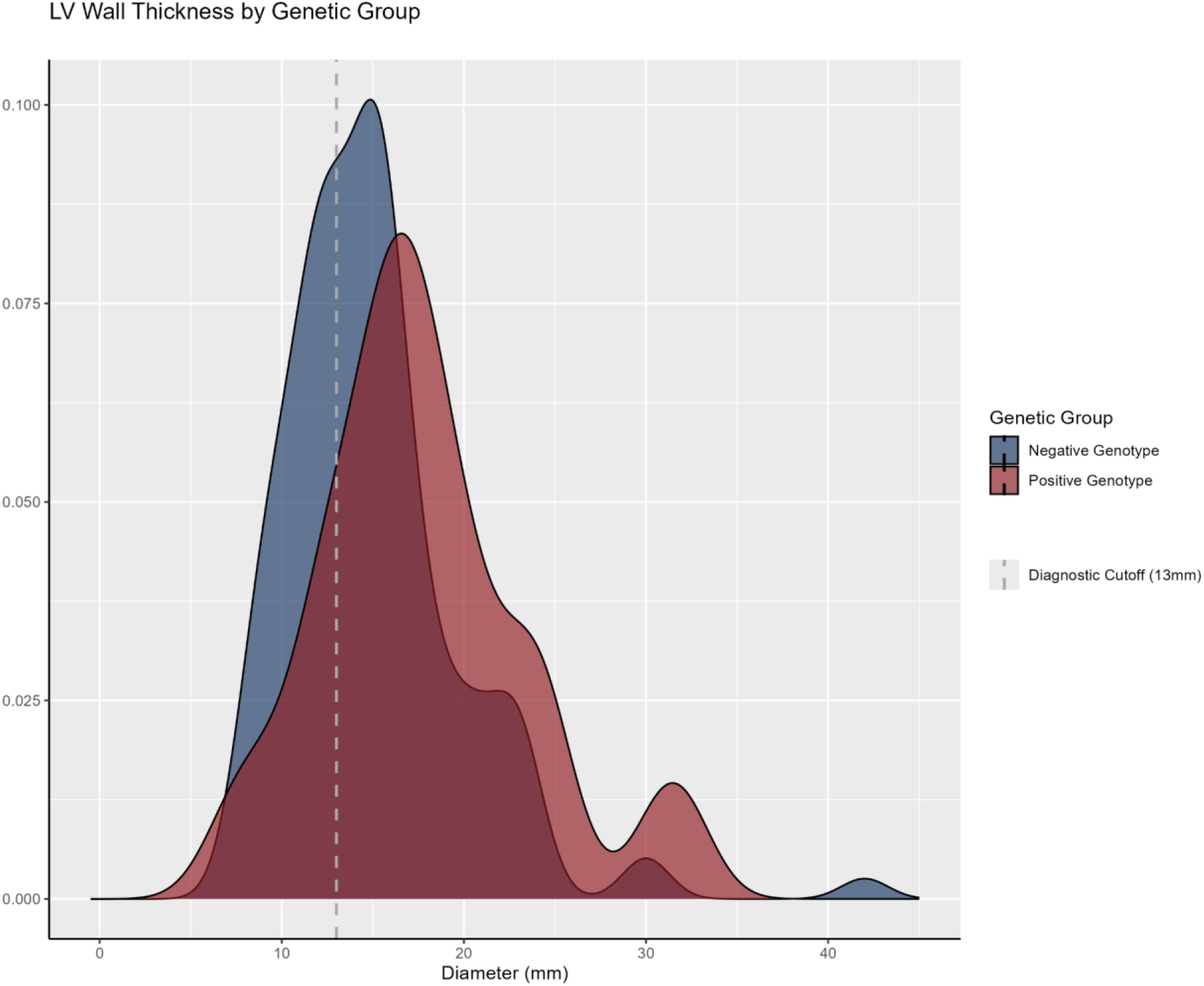
LV Wall by Genetic Group. Shows the distribution of left ventricular wall thickness (measured in millimeters) for genetic diagnosis subgroups: Positive Genotype (Red) and Negative Genotype (Blue). The dashed vertical line at 13mm represents the minimal diagnostic cutoff for HCM based on the 2024 AHA/ACC/AMSSM/HRS/PACES/SCMR guideline.

### Clinical Manifestations and Family History

When it comes to clinical signs and symptoms, palpitations were reported by 61% of the sample, with a higher prevalence in the genotype-positive group (71% vs. 59%; p = 0.042). Dyspnea was observed in 45% of the total sample, with no significant difference between groups (p = 0.5). Syncope occurred in 27% of patients, with similar proportions across groups (p = 0.7). Precordial pain was reported by 44% of participants, again with no significant differences between groups (p = 0.5). Specific manifestations, including angiokeratomas, hypoacusis, and acroparesthesia, also showed no significant variation between groups.

Family history played a pivotal role in characterizing the sample. Parental consanguinity was observed in 11% of cases, while only one individual (0.5%) reported a family history of Fabry Disease. The recurrence of sudden cardiac death among first and/or second degree relatives was reported by 53% of patients diagnosed with HCM, with a higher prevalence in the genotype-positive group (68% vs. 46%; p = 0.004), as illustrated in Table 2.

**Table 2.** Clinical and Demographic Characteristics of the Study Population by Genetic Diagnosis.

|  | Overall<br>N = 200 | Positive Genotype<br>(n = 62) | Negative Genotype<br>(n = 138) | p-value<sup>2</sup> |
| --- | --- | --- | --- | --- |
| **Sample Profile** |  |  |  |  |
| Median Age, y (IQR) | 52 (41 a 66) | 48 (38 a 67) | 54 (43 a 65) | 0.2 |
| Men, n (%) | 115 (58%) | 30 (48%) | 85 (62%) | 0.081 |
| Body mass index, kg/m<sup>2</sup> (IQR) | 27.4 (25.2 a 30.5) | 27.4 (25.5 a 30.7) | 27.3 (24.2 a 30.0) | 0.5 |
| **Signs and Symptoms, n (%)** |  |  |  |  |
| Palpitations | 121 (61%) | 44 (71%) | 77 (56%) | **0.042** |
| Cornea Verticillata | 3 (1.5%) | 2 (3.3%) | 1 (0.7%) | 0.2 |
| Angiokeratomas | 11 (5.5%) | 5 (8.1%) | 6 (4.3%) | 0.3 |
| Dyspnea | 89 (45%) | 30 (48%) | 59 (43%) | 0.5 |
| Syncope | 52 (27%) | 17 (28%) | 35 (26%) | 0.7 |
| Precordial Pain | 87 (44%) | 25 (40%) | 62 (46%) | 0.5 |
| Acroparesthesias | 74 (37%) | 25 (40%) | 49 (36%) | 0.5 |
| **Comorbidity, n (%)** |  |  |  |  |
| Hypertension | 120 (61%) | 34 (55%) | 86 (63%) | 0.3 |
| Diabetes | 40 (20%) | 8 (13%) | 32 (23%) | 0.093 |
| Dyslipidemia | 114 (58%) | 32 (52%) | 82 (60%) | 0.3 |
| ICD Carrier | 9 (5.1%) | 6 (11%) | 3 (2.5%) | **0.026** |
| Hypothyroidism | 17 (9.0%) | 8 (15%) | 9 (6.7%) | 0.10 |
| Heart Failure | 52 (27%) | 18 (31%) | 34 (26%) | 0.5 |
| Renal Failure | 12 (6.6%) | 3 (5.2%) | 9 (7.3%) | 0.8 |
| **Medications, n** |  |  |  |  |
| **(%)** |  |  |  |  |
| Beta-Blockers | 115 (68%) | 42 (78%) | 73 (63%) | 0.054 |
| ACE<br>Inhibitors/ARBs | 88 (52%) | 19 (37%) | 69 (58%) | **0.011** |
| Statins | 101 (59%) | 27 (52%) | 74 (63%) | 0.2 |
| Amiodarone | 17 (11%) | 9 (20%) | 8 (7.3%) | **0.026** |
| Diuretics | 67 (41%) | 20 (41%) | 47 (41%) | >0.9 |
| Aspirin (ASA) | 35 (22%) | 7 (15%) | 28 (25%) | 0.15 |
| **Habits, n (%)** |  |  |  |  |
| Physical Inactivity | 71 (38%) | 23 (38%) | 48 (38%) | >0.9 |
| Smoking | 29 (16%) | 6 (10%) | 23 (18%) | 0.2 |
| Alcohol<br>Consumption | 35 (19%) | 10 (17%) | 25 (20%) | 0.7 |
| **Family History,**<br>**n (%)** |  |  |  |  |
| Parental<br>Consanguinity | 22 (11%) | 4 (6.7%) | 18 (14%) | 0.13 |
| Sudden Cardiac<br>Death | 101 (53%) | 41 (68%) | 60 (46%) | **0.004** |
| **Image-Derived**<br>**Measurements**<br>**, n (IQR)** |  |  |  |  |
| LV Wall Thickness | 15.0 (12.0 a 18.0) | 17.0 (14.2 a 20.6) | 15.0 (12.0 a 17.0) | **<0.001** |
| Ejection Fraction <sup>3</sup> | 66 (61 a 71) | 67 (62 a 72) | 65 (61 a 70) | 0.2 |
| Left Ventricular<br>Diastolic Diameter <sup>3</sup> | 48 (43 a 53) | 46 (42 a 52) | 48 (44 a 55) | **0.030** |
| Left Atrial Diameter <sup>3</sup> | 42 (37 a 48) | 45 (39 a 49) | 41 (36 a 46) | **0.029** |
| 1 Median (Q1 a Q3); n (%) |  |  |  |  |
| 2 Wilcoxon rank sum test; Pearson's Chi-squared test; Fisher's exact test |  |  |  |  |
| 3 Echocardiography |  |  |  |  |

## DISCUSSION

This study evaluated 200 patients through genetic testing for variants in 19 genes associated with hypertrophic cardiomyopathy (HCM) and its phenocopies. The analysis revealed a positivity rate of 31% for pathogenic or likely pathogenic variants, with 24% identified in sarcomeric genes and 7% in genes related to phenocopies. These findings align with medical literature, which reports a diagnostic yield of genetic testing for HCM ranging between 30% and 60% [13,15,16]. A recent multicenter, international study by Silva et al. (2024), utilizing a similar genetic panel, observed a slightly lower positivity rate of 24.7%, with 21.5% attributed to sarcomeric gene variants and 3.2% to phenocopies [17].

The literature commonly cites MYBPC3 as the most prevalent sarcomeric gene, found in 40–45% of cases, followed by MYH7, present in 15–25% of cases. In contrast, our study identified MYH7 and MYBPC3 variants in 46.77% and 14.5% of cases, respectively [3,5,18].

An additional noteworthy observation was the high frequency (6.5%) of variants in the TTR gene within our cohort. While data on the prevalence of TTR variants in the HCM population remain limited, previous studies have reported frequencies ranging from 0.7% to 1.2%. This higher prevalence in our sample may reflect regional genetic variability or a potential founder effect [19,20].

In our study, 89 variants of uncertain significance (VUS) were identified in 81 patients. The medical literature suggests that the penetrance of variants associated with HCM is variable and may be influenced by genetic and environmental factors that are not yet fully understood. Therefore, monitoring these patients, classified in the study as genotype-negative, is crucial due to the possibility of phenotypic conversion over time. This enables the early identification of a transition to a clinical HCM phenotype, allowing for timely therapeutic intervention [3,21].

This study demonstrated that 92,3% of patients, despite originating from different families, with pathogenic or likely pathogenic (P/LP) variants in the TTR gene carried the same variant, Val142Ile, primarily associated with the cardiac phenotype of amyloidosis. Furthermore, these patients had an average age higher than those with sarcomeric gene variants [22]. According to Reddi, the V122I mutation is associated with late-onset disease, frequently observed in individuals of African descent, typically manifesting during the seventh decade of life. Reddi’s study reported a mean disease onset age of approximately 72 years for heterozygotes, whereas in our study, the mean onset age was 70 years [23].

We observed a higher median Maximum Left Ventricular Wall Thickness (MLVWT) in the genotype-positive subgroup compared to the genotype-negative subgroup (17.7 mm vs. mm; p < 0.001). Literature suggests that patients with mutations in sarcomeric genes tend to exhibit greater interventricular wall thickness, often in an asymmetric pattern, with a preference for the apical and anteroseptal regions, and are at increased risk for adverse cardiac events throughout their lifetime. No significant differences were observed between groups or within individual patients in left ventricular measurements obtained through cardiac magnetic resonance imaging (CMR) and echocardiography. This consistency across imaging modalities supports the reliability of these methods for characterizing cardiac morphology in HCM patients [24, 25].

In our study, it was also observed that patients with P/LP variants in sarcomeric genes exhibited a higher incidence of sudden cardiac death (SCD) among first- and/or second-degree relatives. The presence of mutations in sarcomeric genes is associated with an increased risk of SCD [26].

Genetic panels are a relatively recent advancement in medical history and remain largely inaccessible in emerging countries. In this regard, there is a significant portion of the population that may harbor pathogenic or likely pathogenic (P/LP) variants but remain underdiagnosed due to the proband’s premature death, often from sudden cardiac events. This study highlights a higher prevalence of these variants in the examined cohort compared to global prevalence rates, reinforcing the importance of the 2024 AHA/ACC/AMSSM/HRS/PACES/SCMR guideline when advocating for genetic testing in cases with specific clinical indications or under the guidance of a cardiovascular genetics specialist, underscoring its value in improving diagnosis, risk stratification, and familial screening efforts [3,6,11].

When analyzing clinical manifestations, palpitations were more prevalent in the genotype-positive group compared to the genotype-negative group. A systematic review and meta-analysis demonstrated that mutations in sarcomeric genes are associated with an elevated risk of ventricular tachycardia, syncope, and heart failure. Additionally, the greater hypertrophy observed in this group may contribute to a higher prevalence of associated symptoms [26,27].

Analysis of body mass index (BMI) and comorbidities revealed that 28% of patients were classified as obese, 61% had hypertension, 58% had dyslipidemia, and 20% had diabetes mellitus (DM). The literature highlights hypertension as a common comorbidity in patients with HCM, with a prevalence ranging from 35% to 50% in adults. Moreover, studies suggest that patients with coexisting HCM and DM exhibit a higher prevalence of diastolic dysfunction, pulmonary hypertension, and significant mitral regurgitation [28].

Dyslipidemia has been linked to an increased incidence of HCM, particularly in younger individuals, suggesting that dyslipidemia may influence the clinical expression of HCM. Thus, the effective management of these comorbidities is critical to optimizing clinical outcomes and improving the overall prognosis of patients with HCM. [3,29]

The clinical limitations of the present study stem from its design, which was based on the demands of a local population and not composed of a randomized sample of patients. Moreover, being a cross-sectional study, it did not allow for the description of the progression of the pathology in question. Additionally, there is a potential underrepresentation of asymptomatic individuals or those with limited access to specialized care due to the non-random and selective nature of the sample.

The use of advanced technologies, such as cardiac magnetic resonance imaging (CMR) for assessing cardiac morphology, combined with comprehensive genetic analysis, facilitated detailed and precise characterization in this study.

Looking ahead, the future of diagnosing and managing HCM is promising, especially with the continued advancements in genetic testing and imaging technologies like cardiac MRI. This study revealed a higher prevalence of TTR-related cardiac amyloidosis, indicating that regional factors might play a role in genetic variations. Family history and clinical symptoms will become even more important in guiding when genetic testing is needed, leading to earlier diagnoses and more precise risk assessments. With greater access to genetic tests and more personalized treatment options, we can expect significant improvements in the care and outcome for patients with HCM and its related conditions.

The frequency of pathogenic variants in sarcomeric genes aligns with existing literature, but TTR-related cardiac amyloidosis was notably higher, suggesting greater prevalence in this population. Interventricular septal thickness >17 mm and a family history of SCD were strong predictors of positive genetic tests. While genetic testing should be guided by family screening and counseling, a family history of SCD or unconfirmed HCM may justify testing, especially when a pathogenic variant is identified in a family member.

## Data Availability

The code for statistical analysis and the database, in compliance with Brazilian data protection laws, are available at the attached link.

https://github.com/bielkuhn/HCM_Article_UFS

## Nonstandard Abbreviations and Acronyms

ACC: American College of Cardiology
ACEI: Angiotensin-Converting Enzyme Inhibitors
AHA: American Heart Association
AMSSM: American Medical Society for Sports Medicine
ARB: Angiotensin Receptor Blockers
BMI: Body Mass Index
DM: Diabetes Mellitus
ESC: European Society of Cardiology
GLA: Gene associated with Fabry Disease
HCM: Hypertrophic Cardiomyopathy
HRS: Heart Rhythm Society
ICD: Implantable Cardioverter Defibrillator
IQR: Interquartile Range
LVH: Left Ventricular Hypertrophy
MLVWT: Maximum Left Ventricular Wall Thickness
MRI: Magnetic Resonance Imaging
MYBPC3: Myosin Binding Protein C3
MYH7: Myosin Heavy Chain 7
NGS: Next-Generation Sequencing
P/LP: Pathogenic or Likely Pathogenic
PACES: Pediatric and Congenital Electrophysiology Society
RMC: Cardiac Magnetic Resonance
SCD: Sudden Cardiac Death
SCMR: Society for Cardiovascular Magnetic Resonance
TCLE: Termo de Consentimento Livre e Esclarecido (Informed Consent Form)
TTE: Transthoracic Echocardiography
TTR: Transthyretin
VUS: Variants of Uncertain Significance

## ACKNOWLEDGMENTS

The authors acknowledge all investigators and participants who contributed to and made this work possible.

## SUPPLEMENTARY MATERIAL

